# Gut microbiome profiles and associated metabolic pathways in HIV-infected treatment-naïve patients

**DOI:** 10.1101/2020.12.07.20245530

**Authors:** Wellinton M. do Nascimento, Aline Machiavelli, Luiz G. E. Ferreira, Luisa Cruz Silveira, Suwellen S. D. de Azevedo, Gonzalo Bello, Daniel P. Smith, Melissa P. Mezzari, Joseph Petrosino, Rubens Tadeu Delgado Duarte, Carlos R. Zaráte-Bládes, Aguinaldo R. Pinto

**Author notes:** contributed equally to this work (Carlos R. Zaráte-Bládes, Aguinaldo R. Pinto).

## Abstract

The normal composition of the intestinal microbiota is a key factor for maintaining health homeostasis and, accordingly, dysbiosis is well known to be present in HIV-1 patients. Here, we investigate the gut microbiota profile of HIV-1 positive patients without antiretroviral therapy and healthy donors living in Latin America. We enrolled 13 HIV positive patients (six elite controllers, EC and seven non-controllers, NC) and nine healthy donors (HD). Microbiota compositions in stool samples were determined by sequencing the V3-V4 region of the bacterial 16S rRNA and functional prediction was inferred using PICRUSt. Several taxa were enriched in EC compared to NC or HD groups, including *Acidaminococcus, Clostridium methylpentosum, Barnesiella, Eubacterium coprostanoligenes* and *Lachnospiraceae UCG-004*. Importantly, we confirmed that the route of infection is a strong factor associated with changes in gut microbiome composition and we extended these results by identifying several metabolic pathways associated with each route of infection. Moreover, we observed several bacterial taxa associated with different viral subtypes such as *Succinivibrio* which were more abundant in patients infected by HIV subtype B, and *Streptococcus* enrichment in patients infected by subtype C. In conclusion, our data brings a significant contribution to our understanding of dysbiosis-associated changes in HIV infection and describes, for the first time, differences in microbiota composition according to HIV subtypes.

## 1. INTRODUCTION

Human immunodeficiency virus 1 (HIV-1) infection is estimated to affect 38 million people worldwide [1]. Chronic HIV infection is characterized by substantial depletion of CD4^+^ T cells, followed by disruption of intestinal epithelial barrier, gut microbiota dysbiosis, microbial translocation, and chronic activation of the immune system [2]. As HIV-1 replicates since early infection in gut associated lymphoid tissue (GALT), it is considered a mucosal disease with systemic manifestations.

Dysbiosis has been well documented in HIV-1 patients, [3,4] and studying the interplay between the microbiome and HIV-1 infection is pivotal in order to better understand the disease and to identify novel approaches which can improve treatment [2]. Nonetheless, studies evaluating the composition of microbiota in HIV-1 infected individuals show great variability and some conflicting results, making it difficult to identify specific bacterial species, genera or even families associated to specific subgroup of HIV patients in different populations [5–7]. This is in part due to the fact that the dysbiosis profile in HIV-1 infection varies substantially among individuals across the globe, which stresses the necessity of microbiome evaluations to be performed in different geographical regions to validate shifts in microbiome composition and/or to identify which bacterial components are equivalent to those observed in HIV-1 patients from other regions [8,9].

In this study we performed the first microbiota analysis on HIV-1 patients and healthy controls of individuals both from and living in Latin America. We confirmed several differences in microbiota composition of HIV-1 infected individuals according to RNA viral load levels and the route of infection, and we show for the first time that patients display a distinct microbiota profile according to HIV-1 subtype. The results provide opportunities for the identification of new molecular targets in the development of complementary therapeutics against HIV infection.

## 2. METHODS

### 2.1 Study population

This study was conducted with a cohort of 13 individuals with documented HIV-1 infection (HIV+) for over five years and who maintain CD4^+^ T cell counts >500 cells/mm^3^ and RNA viral loads <20,000 copies/mL without antiretroviral therapy (ART). These subjects were classified in two groups: (i) elite controllers (EC) with (100%) plasma viral load (VL) determinations undetectable (<50 copies/mL, n = 6), and (ii) HIV non-controllers (NC) with most (≥70%) VL determinations between 2,000 and 20,000 copies/mL (n = 7). We also included nine healthy donors (HD) as controls. Volunteers were recruited at Hospital Regional Homero de Miranda Gomes, Hospital Nereu Ramos and at Blood Bank from the Hospital Universitário, Universidade Federal de Santa Catarina (UFSC), all located in the greater Florianópolis area, Santa Catarina, Brazil. The UFSC Ethics Committee (Comitê de Ética em Pesquisa com Seres Humanos, CEPSH-UFSC) approved the study under protocol number 1.622.458, and all volunteers provided written informed consent according to the guidelines of the Brazilian Ministry of Health.

### 2.2 Sample collection and processing

Stool samples were collected in a sterile plastic container either at the volunteers’ residence or at the hospital. DNA was extracted using the commercial kit QIAamp DNA Stool Mini Kit (Qiagen, Germany) and stored at −20°C. As negative control, a sample of sterile water was used. Data on CD4^+^ and CD8^+^ T cells count and plasma HIV-1 VL were accessed via medical records and measured according to the Brazilian Ministry of Health guidelines [10].

### 2.3 Viral subtype analysis

To identify the viral subtype, total DNA was extracted from peripheral blood mononuclear cells (PBMCs) and the HIV-1 *env* gene was amplified by nested PCR and sequenced as previously described [11]. The chromatograms were assembled using SeqMan 7.0 software (DNASTAR Inc., USA) and consensus e*nv* sequences covering positions 7,008–7,650 relative to the HXB2 reference genome were generated. The *env* sequences were aligned with subtype reference sequences downloaded from the Los Alamos HIV Sequence Database (http://www.hiv.lanl.gov/) using ClustalW and then manually edited and subtyped by maximum-likelihood (ML) phylogenetic tree reconstructions with the PhyML 3.0 program as described previously [11].

### 2.4 Sequencing and bioinformatics analysis

DNA was quantified using the Qubit dsDNA HS kit (Thermo Fisher Scientific, USA). The targeted V3-V4 region of the bacterial 16S rRNA gene from the extracted DNA was PCR-amplified using the Illumina primers S-D-Bact-0341-b-S-17 (forward) and S-D-Bact-0785-a-A-21 (reverse) [12]. The polymerase chain reactions were performed as described by Machiavelli *et al*. [13] DNA sequencing was performed using a V2 reagent kit 2 × 250 bp (500 cycles) (Illumina, MS-102–2003) at the Illumina MiSeq platform. Raw reads were quality filtered using Trimmomatic v0.38 [14] according to sequence size and Phred score. The three first nucleotides in all sequences presented a Phred score below 20 and were removed. Sequences with less than 200 or more than 259 nucleotides were also excluded. Paired-end joining, determination of amplicon sequence variants (ASVs), and removal of chimeric sequences were performed using the DADA2 [15] R package v1.15.5. Taxonomy was assigned with the DECIPHER R package v2.10.2 IDTAXA [16] algorithm and the SILVA_SSU_r138_2019 [17]. Finally, the assigned taxonomy was organized in a BIOM-formatted ASV table and imported into R and ATIMA (Agile Toolkit for Incisive Microbial Analyses) software for statistical analysis. Functional prediction was performed using PICRUST v2.1.4-b, [18] based on the Kyoto Encyclopedia of Genes and Genomes (KEGG). Counts were normalized by considering 16S rRNA gene copy number. The sequencing data was deposited in the NCBI Sequence Read Archive.

### 2.5 Statistical analysis

Statistical analyses were performed using R software v3.6.0 and ATIMA v3.0. Characteristics between groups were compared using the Kruskal-Wallis and Mann-Whitney test. Gut microbiome taxa abundance was assessed using the phyloseq R package and alpha diversity indices were assessed using ATIMA software. Kruskal-wallis and Mann–Whitney test were applied to compare the differences in the number of observed ASVs, alpha diversity richness (Chao1), and diversity (Shannon and Simpson) by groups. Beta diversity was evaluated using the Principal Coordinate Analysis (PCoA) on Bray-Curtis, weighted Unifrac and unweighted Unifrac distances to determine significant differences between groups. Linear discriminant analysis (LDA) effect size (LEfSe) [19] algorithm was used to identify differentially abundant bacterial taxa between the study groups, using the online version of Galaxy. LDA was performed using one-against-all strategy and score of >2.0. LEfSe uses the Kruskal-Wallis and Wilcoxon rank-sum test to detect features with significantly different abundances between assigned taxa and LDA to estimate the effect size of each feature. Functional prediction was analyzed using ATIMA software applying the Mann–Whitney test to compare the relative abundance between groups. *P* values below 0.05 were considered significant after multiple-comparison correction using the false discovery rate (FDR) method.

## 3. RESULTS

### 3.1 Epidemiological features

This was a cross-sectional study and detailed information regarding demographic and clinical characteristics of participants (13 HIV+ and nine healthy donors, HD) is displayed at Table 1. HIV+ individuals were separated in two groups: elite controllers (EC, n = 6) and non-controllers (NC, n = 7) according to the plasma VL. All EC showed undetectable plasma viral load without blips at all measurements while the median level of plasma viremia for NC was 4,203 copies/mL. EC and NC had documented HIV-1 infection for a median of 5.6 (IQR 5.0-9.3) years and 6.4 (IQR 5.8-6.9) years, respectively. The median of CD4^+^ T cell counts in EC and NC was 1,190 (IQR 605.5- 1,370) and 726 (IQR 521-866) cells/µL, respectively. As for CD8^+^ T cells, the median was 1,057 (IQR 810-1,356) cells/μL for EC and 1,295 (IQR 798-1,527) cells/μL for NC. Phylogenetic analysis using *env* sequences showed that our cohort included two HIV-1 subtypes: B (n=6, 46%) and C (n=7, 54%). Four EC, one NC and one HD declared use of antibiotics within three months before enrollment in the study.

**Table 1.**
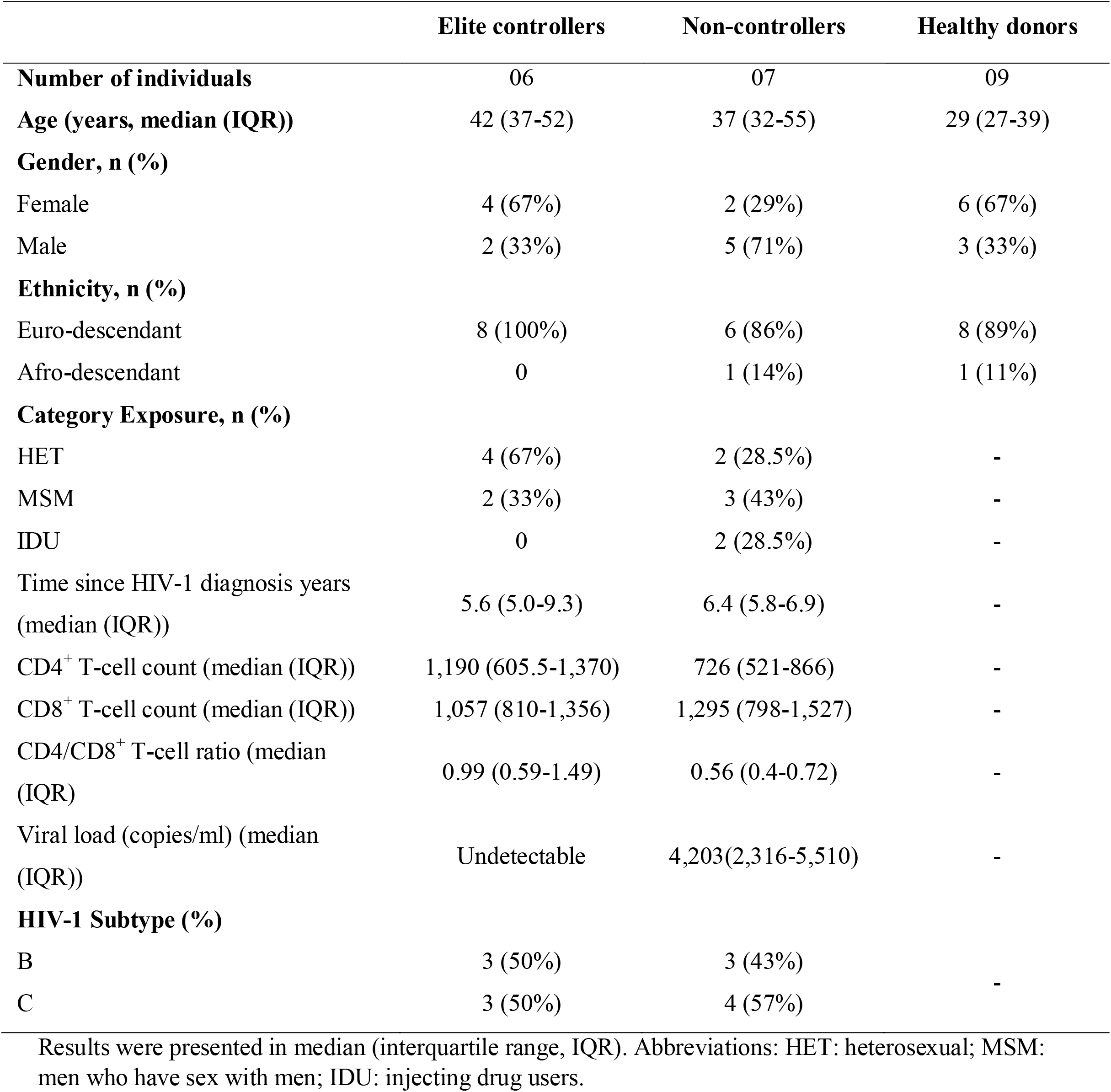
Demographic and clinical characteristics of participants.

### 3.2 Comparison of microbiota profile between HIV+ and HD

The gut microbiota characterization was analyzed using 16S rRNA sequencing of stool. A total of 2,399,610 reads with an average of 99,983 reads per sample were generated after data processing and quality check, resulting in a final count of 1.001 ASVs. In terms of microbial composition, 14 main phyla were found, regardless of the HIV status. The Firmicutes, Bacteroidota, Proteobacteria and Actinobacteriota phyla were the most abundant (Fig. 1A). There was no statistical difference between HIV+ and HD in beta and alpha diversity (Fig S1). To determine and distinguish differentially abundant ASVs associated with HIV-1, we compared the gut microbiota composition between HIV+ and HD using the LEfSe algorithm, which showed significant bacterial differences between HIV+ and HD. The family Atopobiaceae, and genera *Acidaminococcus, Libanicoccus, Lachnospiraceae NK3A20 group* were enriched in HIV+, whereas the order Verrucomicrobiales, class Verrucomicrobiae, families Barnesiellaceae and Akkermansiaceae and genera *Angelakissela, Oscillospira, Turicibacter, Bilophila, Barnesiella, Akkermansia* were enriched in HD (Figure 1B, C). The relative abundance of taxa identified in LEfSe as differentially abundant between HIV+ and HD was compared using the Mann-Whitney test, which revealed statistically significant differences between groups in seven of the nine identified genera (*P*< 0.05) (Fig 1D).

**Figure 1.**
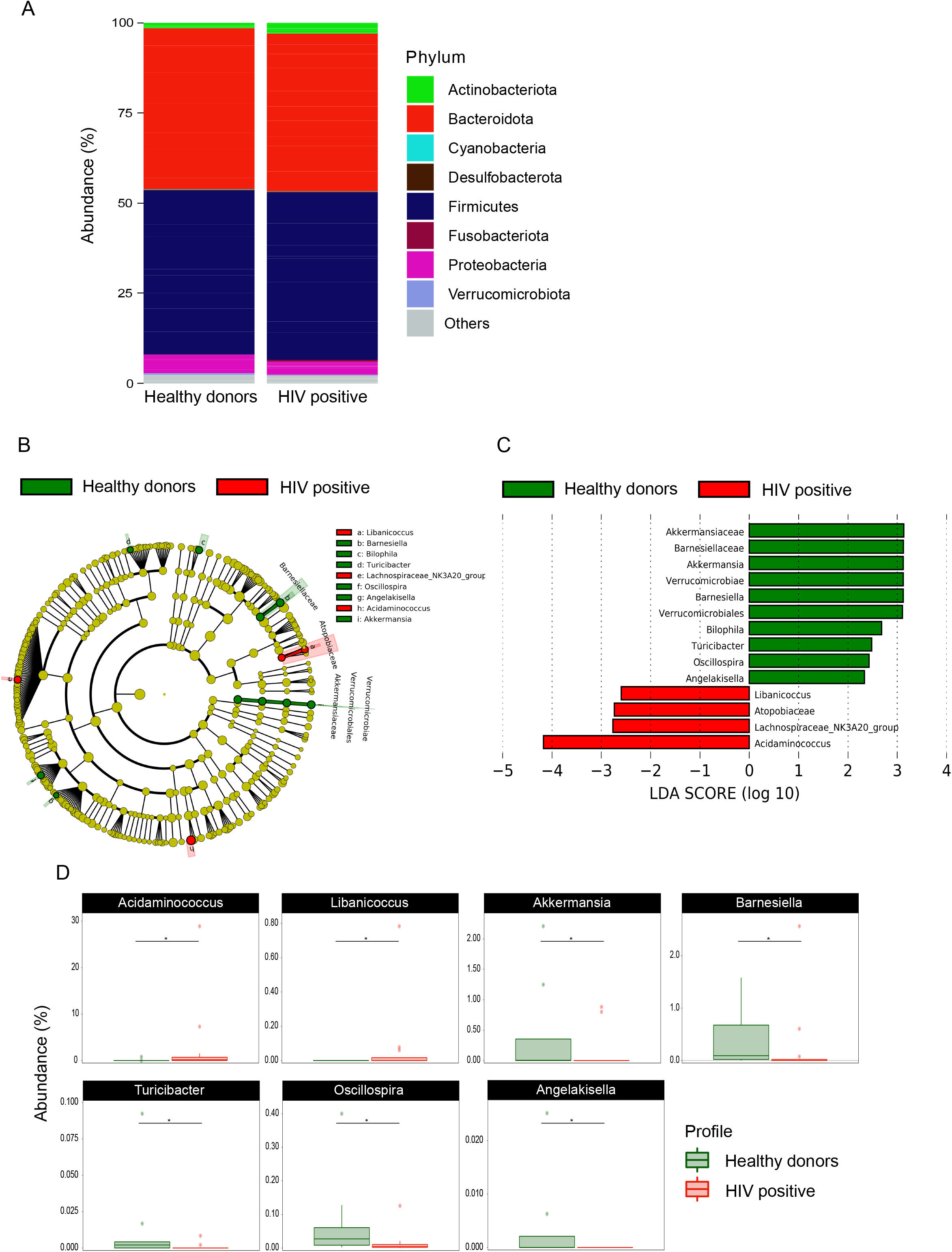
Taxonomic differences in fecal microbiota between HIV-1 positive patients and healthy donors. Relative abundance at phylum level (a); Cladogram of LEfSe linear discriminant analysis (LDA) scores showing differentially abundant taxonomic clades with an LDA score >2.0 in gut microbiota of HIV-1 patients and healthy donors (b); LDA scores of differentially abundant taxa in gut microbiota of HIV-1 patients and healthy donors (c). The relative abundance of genera identified by LEfSe as being differentially abundant in HIV-1 patients and healthy donors were compared using the Mann–Whitney test (d). \**P* <0.05.

### 3.3 Differences in microbiota composition between EC, NC, and HD

LEfSe analysis was used to compare the gut microbiota of EC, NC and HD groups. Figure 2A and B show differences in the abundance of taxonomic clades of LDA score >2.0 among EC, NC and HD individuals. The gut microbiota of the EC group was dominated by taxa *Acidaminococcus, Clostridium methylpentosum group, Barnesiella, Eubacterium coprostanoligenes group* and *Lachnospiraceae UCG-004*, whereas the NC group showed high scores for the family Atopobiaceae, and genera *Libanicoccus* and *Lachnospiraceae NK3A20 group*. The features identified by LEfSe as being differentially abundant were confirmed by standard statistical analysis using Kruskal-Wallis test (Fig. S2). PICRUSt analysis was performed to determine whether the taxonomic differences between the groups’ microbiota corresponded to functional changes. We found no significant differences between HD and EC groups or between EC and NC groups (data not shown). However, we identified 30 KEGG pathways as significantly different between NC and HD groups (Fig. 2C). PICRUSt analyses demonstrated that the NC group’s microbiota exhibits increased S-adenosyl-L-methionine cycle, methylerythritol phosphate pathway, and biosynthesis of threonine, acetylmuramoyl pentapeptide, peptidoglycan, CoA, and CDP-diacylglycerol. Conversely, the microbiota of NC subjects were depleted for sucrose and glycogen degradation, biosynthesis of thiamine, folate, fatty acids, unsaturated fatty acids and lipopolysaccharides, B12 Salvage from cobinamide and sulfur metabolism (*P* < 0.05).

**Figure 2.**
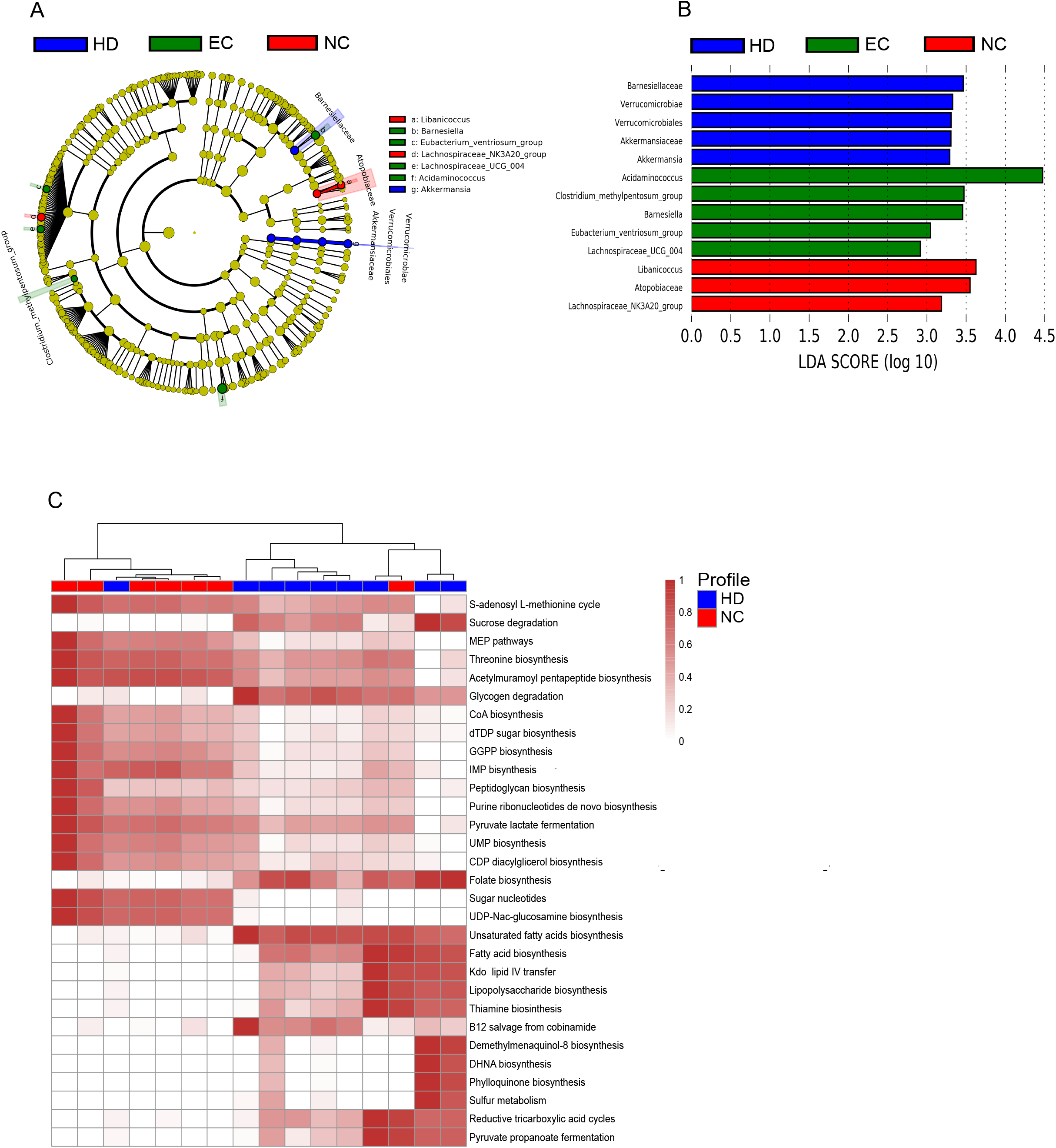
Taxonomic differences and inferred functional content of gut microbiota of healthy donors (HD), elite controllers (EC) and non-controllers (NC). LEfSe results for the bacterial taxa that were significantly different between HD, EC and NC groups. Cladogram showing differentially abundant taxonomic clades (a) and LDA scores showing significant differences between groups (b). Comparison of PICRUSt predicted KEGG function. The heatmap shows the relative abundance of pathways that were significantly different between HD and NC groups (c). Significant differences between groups were tested with Mann-Whitney test (FDR-adjusted *P*-value <0.05).

### 3.4 HIV transmission route influences composition of gut microbiome

HIV+ patients were divided into three groups according to exposure category: HET (n= 6), MSM (n= 5) and IDU (n= 2). Beta diversity with Principal Coordinates Analysis of Bray-Curtis distances indicated differences in the gut microbiota between HET, MSM and IDU groups, (*R* = 0.36, *P* = 0.011) (Fig. 3A). We carried out LEfSe analysis to determine the taxa most associated with the route transmission of HIV-1 and significant differences in the abundance of 27 taxa among HET, MSM and IDU groups were found. Bacteroidaceae, *Bacteroides, Alistipes, Holdemania, Unclassified lactobacillales, Subdoligranulum, DTU089, UBA1819, Unclassified Clostridium methylpentosum group, Clostridium methylpentosum group, Eisenbergiella* and *Odoribacter* were enriched in faeces of HET group, whereas Aeromonodales, Succinivibrionaaceae, VadinBE97, *Prevotella, Succinivibrio, Megasphaera, Unclassified vadinBE97, Allisonella, Libanicoccus* and *Mitsuokella* were enriched in MSM group (Fig 3B and C). When differences in microbiota functions were assessed, 20 pathways strengthened and 30 pathways weakened in MSM in relation to the HET group (Fig 3D). Alanine biosynthesis, metabolic regulators, sulfur oxidation and biosynthesis of arginine carbohydrates, biotin, fatty acid, and tryptophan were among the enriched pathways in MSM group compared to the HET group, while biosynthesis of valine, histidine, phospholipid, acetylmuramoyl pentapeptide as well as purine, glycogen and sucrose degradation were depleted (*P* < 0.05).

**Figure 3.**
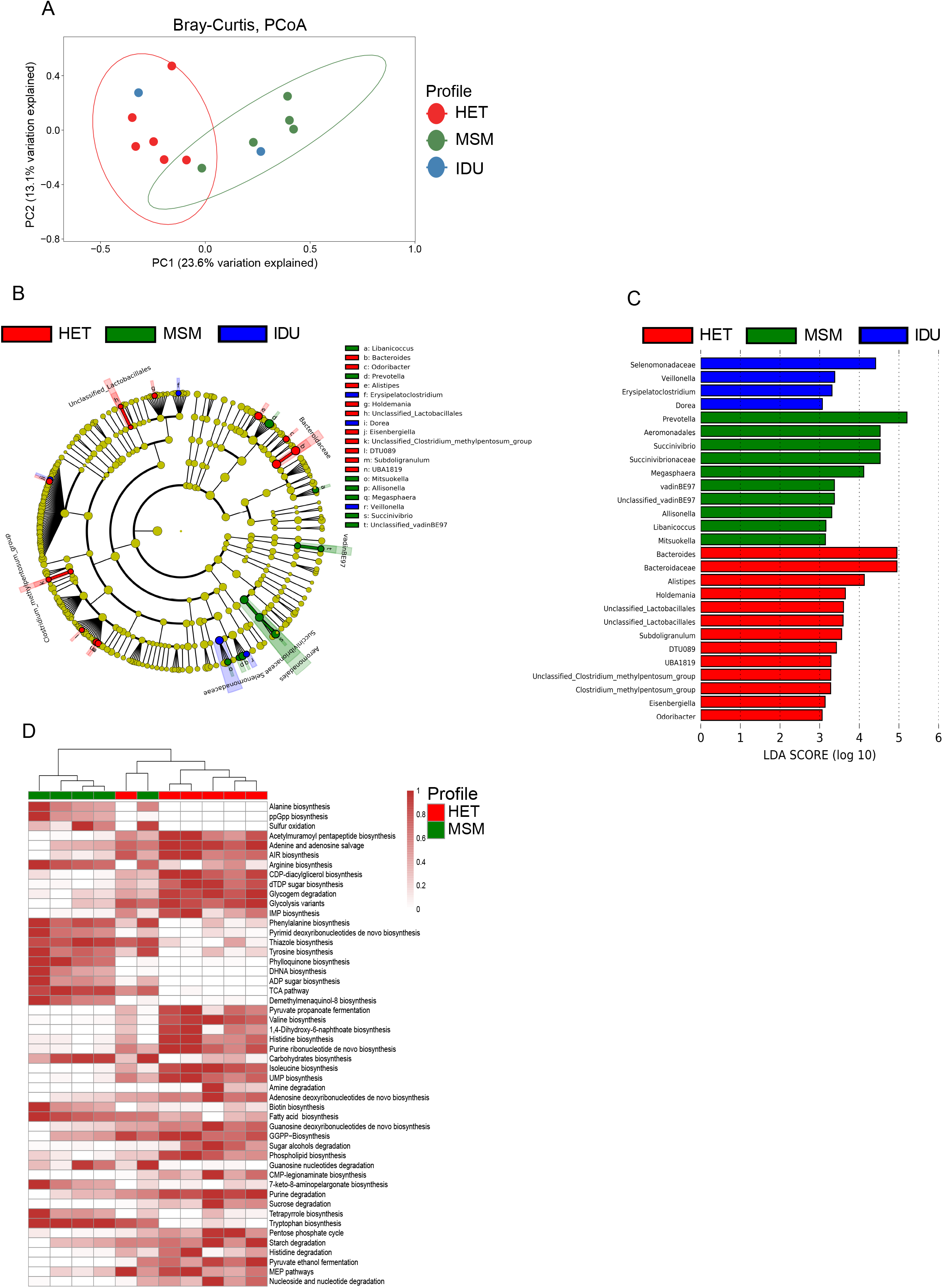
Effects of HIV transmission route on gut microbiota of HIV-1 positive patients. Principal Coordinate Analysis (PCoA) of Bray Curtis distances among Heterosexual (HET), men who have sex with men (MSM) and injecting drug users (IDU) groups (a). LEfSe results for the bacterial taxa that were significantly different between HET, MSM and IDU groups. Cladogram shows differentially abundant taxonomic clades (b), and LDA scores showing significant differences between groups (c). Heatmap of relative abundance pathways that were significantly different between HET and MSM groups (d). Significant differences between two groups were tested with Mann-Whitney test (FDR-adjusted *P*-value <0.05).

### 3.5 Gut bacterial microbiota profile differs between patients infected with HIV-1 subtype B and C

Next, we explored whether changes in gut microbiota occur according to HIV-1 subtype and found clear differences in the composition of gut microbiome among patients infected with subtype B (HIV-1B) or C (HIV-1C). Figure 4A and B show that abundance of Lentisphaeria class, Aeromonadales, Victivallales order, Succinivibrionaceae and VadinBE97 family, and *Succinivibrio, Megasphaera, Prevotellaceae UCG-003, Mitsuokella, Solobacterium, Unclassified vadinBE97, Asteroleplasma, Parvimonas, Unclassified Parvimonas, Anaerovibrio* and *Alissonela* genus were more abundant in HIV-1B group, whereas Lactobacillales order, Selenomonadaceae, Streptococcaceae and Enterococcaceae family, *Streptococcus, Atopobium, Eggerthella, Veillonella, Christensenella* and *Enterococcus* genus were more abundant in HIV-1C group. Differences in bacterial abundance at genus level between these two groups can be seen at Figures 4C and D.

**Figure 4.**
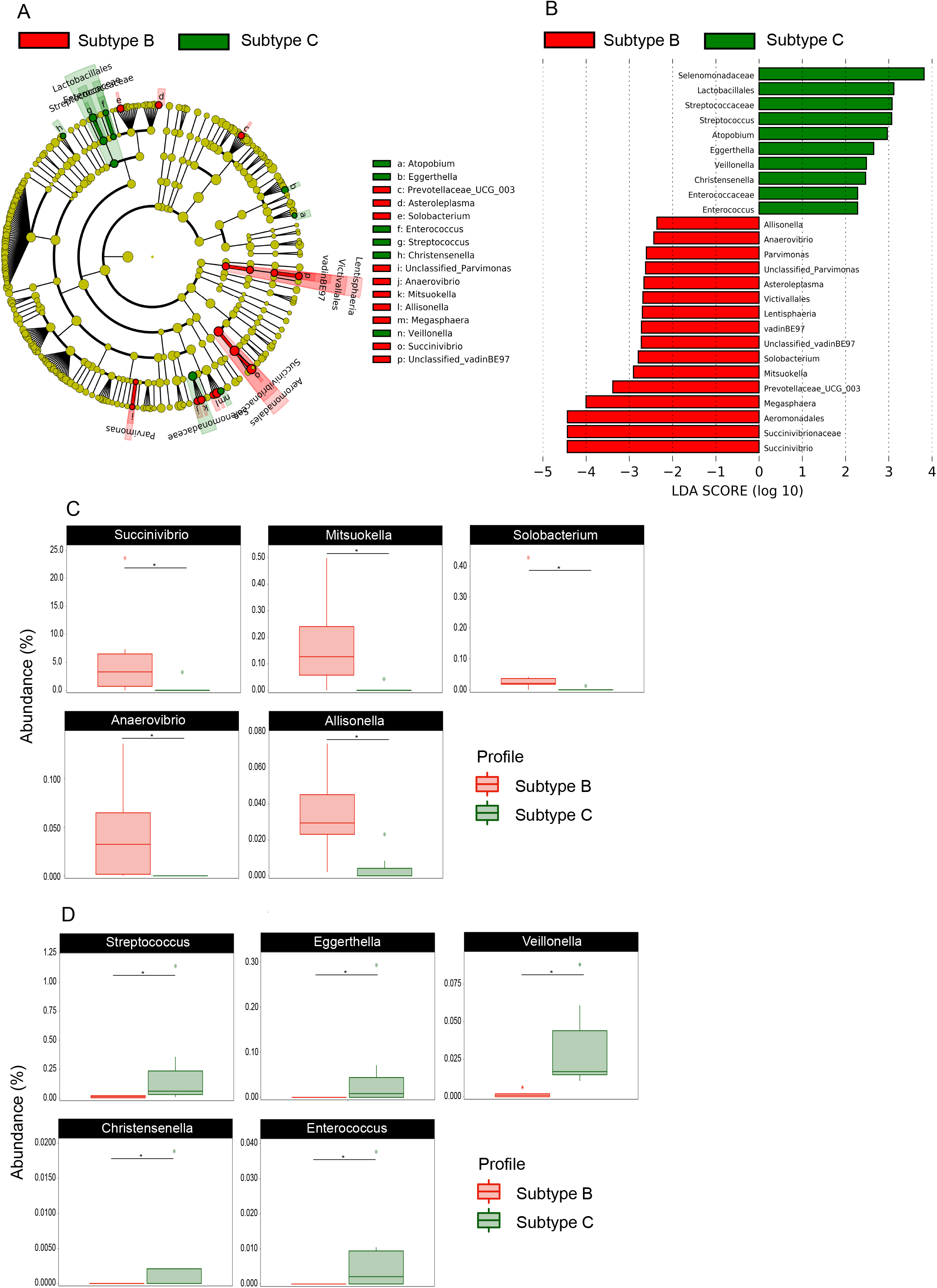
Effects on gut microbiota according to HIV subtype B and C. LEfSe results for the bacterial taxa that were significantly different between HIV positive patients infected by subtype B or C. Cladogram shows differentially abundant taxonomic clades (a) and LDA scores showing significant differences between groups (b). The relative abundance of genus identified by LEfSe as being differentially more abundant in HIV positive patients infected by HIV subtype B (c) or subtype C (d) were compared using the Mann–Whitney test (c). \**P* values <0.05.

## 4. DISCUSSION

Our study is the first to analyze gut microbiota composition in HIV-1 infected patients both from and living in Latin America (more specifically in Brazil), including a group of EC individuals. Along with other reports, [5,6] our data did not reveal differences in microbiota alpha and beta diversity between HIV+ and HD. This finding contrasts with other studies where such differences have been identified [8,9]. Together, these data indicate not only a lack of consensus about differences in alpha and beta diversity in HIV+ in relation to HD, but also that microbiome alterations in HIV+ may vary across populations, likely due to factors that have not always been considered, such as genetic background, different types of nutritional habits, ART, the clinical differences among HIV+ patients and even HIV subtype.

The deeper analyses performed in this study revealed a number of important differences. The HIV+ gut microbiota were enriched with *Acidaminococcus* genus, similar to a previous report [20]. *Akkermansia* genus was also enriched in HIV+ individuals, in agreement with Mutlu *et al*.[8] findings. *Akkermansia* is a gram-negative anaerobic genus abundantly found in the human gastrointestinal tract throughout life, [21] with several proposed roles such as mucin degradation, [21,22] anti-inflammatory activity, [23] immunomodulation properties on the adaptive immune system [24] and protective barrier functions [25]. Some other microorganisms were less abundant in the gut of HIV+ individuals, such as members of Lachnospiraceae family which have been implicated in butyric acid production [26] that is associated to protection of colon cancer, [27] influences obesity levels, [28] as well as assists maintaining the epithelial barrier [29].

We found that the EC group presented a unique signature composed by *Acidaminococcus, Barnesiella, Clostridium methylpentosum group, Eubacterium ventriosum group* and *Lachnospiraceae* UCG*-004. Barnesiella* genus, in particular, has been associated with beneficial effects on human gastrointestinal tract [30]. Interestingly, although several previous studies have reported members of the Lachnospiraceae family associated with HD, [8] here we found an increased abundance of *Lachnospiraceae UCG-004* in EC patients. It is tempting, based on our findings, to speculate that this genus could be considered as a marker of HIV-1 control in EC patients.

We also identified differences in metabolic pathways that varied in abundance between NC and HD groups, such as pathways involved in carbohydrate and lipid metabolism, which were reduced in NC individuals. It is known that specific changes in metabolic pathways are associated with effector functions of the immune system. For example, it has been suggested that the synthesis of fatty acids is involved in the induction of inflammatory responses of macrophages [31]. Vesterbacka *et al*. [7] also found pathways involved in carbohydrate metabolism that was significantly reduced in HIV+ ART-naive patients compared to HD. This same study also demonstrated that some pathways related to lipid metabolism, including fatty acid metabolism and lipid biosynthesis proteins were less abundant in HIV+ ART-naive patients, while linoleic acid metabolism was more represented in this same group. We also detected pathways related to metabolism of cofactors and vitamins, energy metabolism, and glycan biosynthesis that were less represented in NC patients compared to HD subjects.

Similar to what has been observed by Noguera-Julian *et al*. [9] the microbiome profile of our patients clustered according to HIV transmission group, in which HET and MSM microbiomes behave distinctly, whereas the IDU group did not cluster separately, although it displayed some taxa differences compared to the other two groups. Remarkably, *Prevotella* and *Bacteroides* were associated to MSM and HET groups, respectively, as previously reported in individuals from Barcelona and Stockholm.[9] Moreover, we also identified an association between *Succinivibrio* in the MSM group and between *Alistipes* and the HET group. In addition to the previously reported anti-inflammatory capacity, [32] an increased abundance of the *Succinivibrio* genus has been found in patients with cervical cancer in comparison to HD, [33] whereas *Alistipes* genus is an anaerobic bacteria found commonly in the healthy human gastrointestinal tract [34].

Functional analysis of microbiome associated to the transmission route also identified a high number of significantly abundant pathways in HET and MSM, extending the important observations of a previous study [9] and confirming that sexual preference is a strong factor associated with gut microbiota composition in HIV-1+ patients. For example, pathways involved in amino acid metabolism were differentially distributed between HET and MSM groups. It has been shown that the metabolism of various amino acids can play an important role in mediating the functionality of immune cells [31]. Pathways such as histidine degradation, isoleucine, valine and histidine biosynthesis were enriched in HET groups, while that alanine, phenylalanine tyrosine and tryptophan biosynthesis were more abundant in MSM group. Additionally, we identified a lower abundance of pathways involved in carbohydrate metabolism in HET group. In contrast, some pathways related to metabolism of cofactors and vitamins were more enriched in MSM group. Although these results contribute in the understanding of HIV-1 infection, it would be important to identify which of these metabolic connections are established and truly relevant for the disease history so that it can be identified whether gut bacterial metabolism is altered as a consequence of sexual preference. This could potentially be used to target certain molecular pathways as a complementary treatment option to improve patients’ quality of life by reducing the impact of gut microbiota dysbiosis.

Finally, the main finding of our study is that HIV+ patients had a distinct microbiota according to viral subtype B or C. High molecular variability is a striking feature of HIV-1 in the global AIDS pandemic, which has evolved into a multiple number of variants [35]. Although HIV-1B dominates in many countries in Europe and the Americas, more than 50% of the infections worldwide are caused by HIV-1C, which is the most prevalent subtype in southern African countries and India [36]. In the southern region of Brazil, however, HIV-1B and HIV-1C co-circulate, making this region a unique place for epidemiological studies [37]. The importance of subtypes in different clinical parameters of HIV-1 infection is still controversial, but the successful worldwide dissemination of subtype C may be due to this strain being less virulent in comparison with other subtypes while maintaining the same transmission efficiency [38]. In a study conducted by Venner *et al*. [39] in which women from the same area and infected by subtypes A, C or D were followed for 9.5 years and disease progression was evaluated, it was mentioned that it would be important to further evaluate the vaginal microbiota as a way to access disease progression differences according to viral subtype within that cohort.

As far as we know, the present report is the first to associate microbiota composition with HIV subtypes. Dang *et al*. [40] indirectly associated higher colonization of *Streptococcus* species in oral microbiota of HIV-1 patients on ART when compared to HD and ART naive HIV+ patients, all infected by HIV-1 subtype B. Our results showed *Succinivibrio, Mitsuokella, Solobacterium, Anaerovibrio* and *Allisonella* associated to HIV-1B, while *Streptococcus, Eggerthella, Veillonella, Christensenella* and *Enterococcus* genus were more abundant in individuals infected by subtype C. Taken together, these data highlight the importance of our findings regarding the difference in gut microbiota profile according to HIV-1B and HIV-1C infection. Although HIV-1 does not target bacteria, it is worth considering what effects viral subtypes could exert on the immune system and subsequently impact on gut microbiota. Moreover, our results open the possibility to consider targeting microbiota as a way to improve the response to HIV-1 infection through the effects of viral subtypes on dysbiosis.

We acknowledge that while our study has the ability to show associations, we can only suggest causality to a limited extent. In addition, our study has an important limitation due to the low number of participants. Nonetheless, our cohort included only ART naïve HIV+ patients diagnosed at least 5 years before the time of recruitment and showed for the first time an association of HIV viral subtype B and C with distinct gut microbiota profile.

## Data Availability

The authors authorize the availability of all data mentioned in the study.

## Author statement

WM: designed the study, recruited volunteers, processed samples, analyzed the data, and wrote the manuscript. AM and LCS analyzed the data. LGEF: designed the study and recruited volunteers. SSDA, GB, DPS, MPM, JP and RTDD helped with data analysis and revised the manuscript. ARP and CRZB designed the study, supervised the work and revised the manuscript. All authors read and approved the final version of the manuscript.

## Acknowledgments

The authors would like to thank all the volunteers who participated in this study and the team at the Regional Hospital Homero de Miranda Gomes and Blood Bank from the Hospital Universitário/UFSC who assisted in the recruitment of volunteers and collection of samples. The authors also thank Dr. Oscar Bruna-Romero for providing reagents.

---

**Figure S1.**
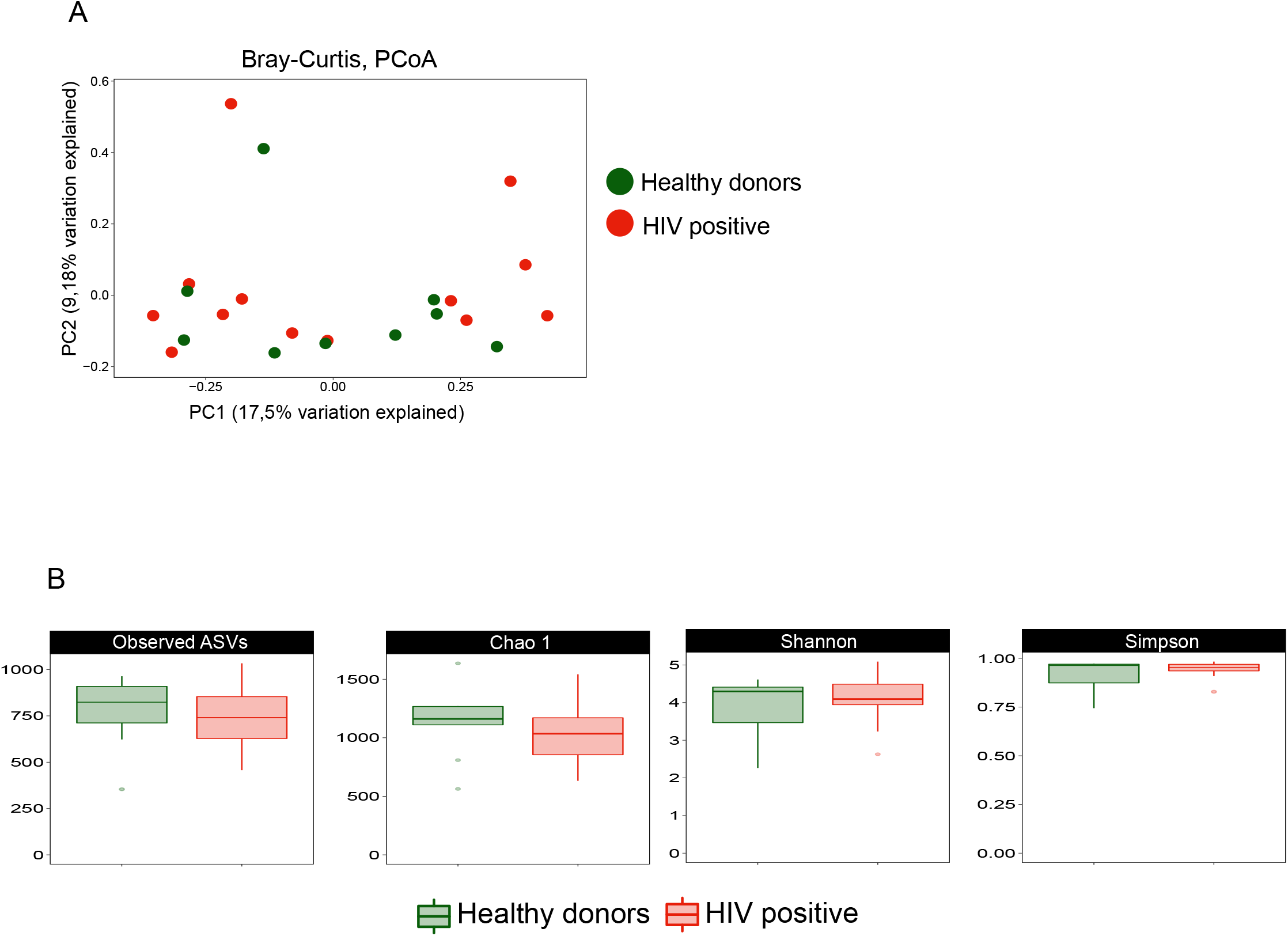
Comparison of gut microbiota of HIV-1 positive and healthy donors. PCoA of Bray Curtis distances among groups (a). Total number of ASVs observed, Richness (Chao1) and diversity (Shannon and Simpson) indices (b).

**Figure S2.**
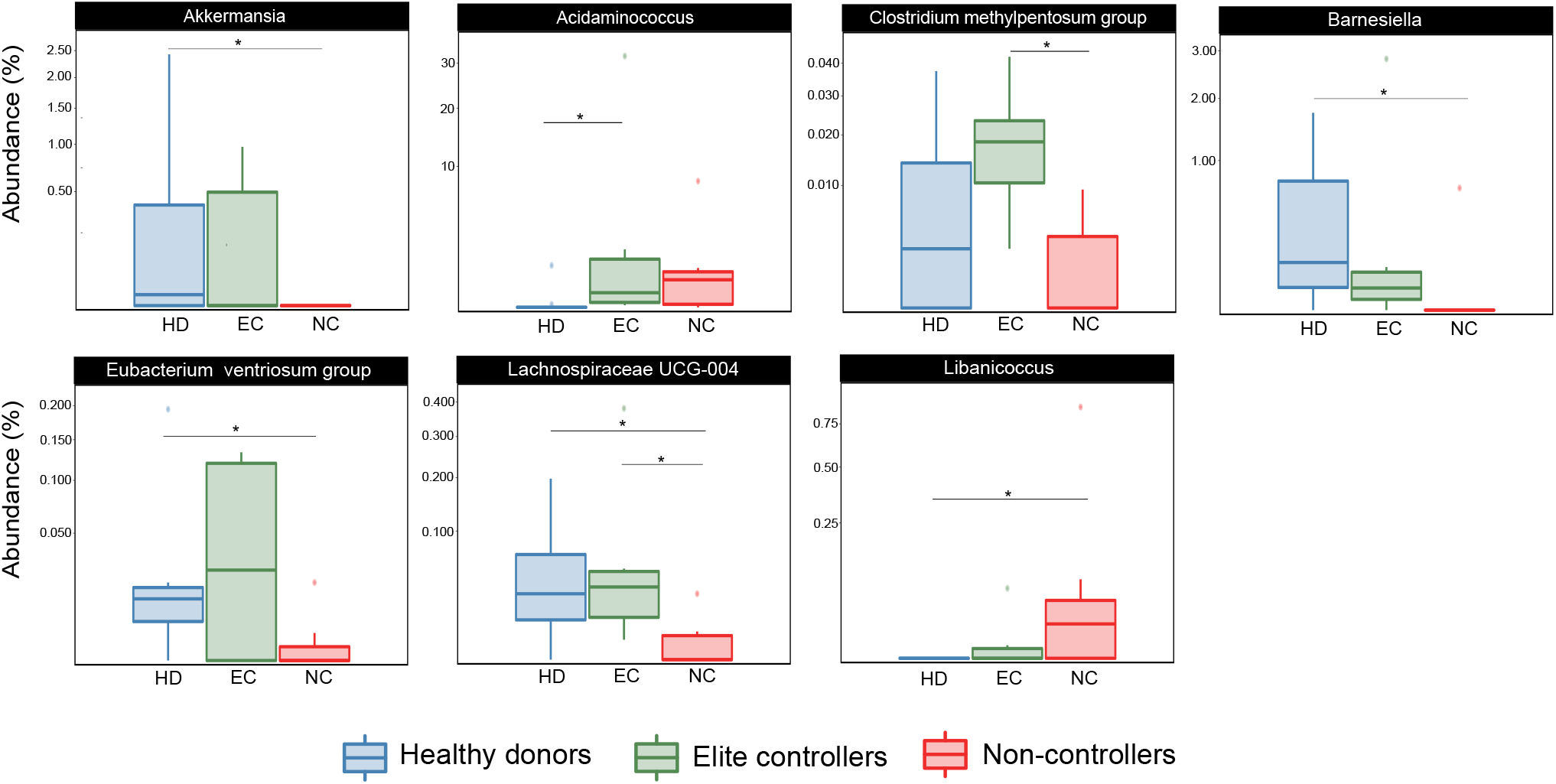
Comparison of gut microbiota of healthy donors (HD), elite controllers (EC) and Non-controllers (NC). The relative abundance of genera identified by LEfSe as being differentially abundant between groups were compared using the Kruskal-Wallis test. \**P* values <0.05.

